# Hybrid and vaccine-induced immunity against SARS-CoV-2 in MS patients on different disease-modifying therapies

**DOI:** 10.1101/2022.06.28.22276989

**Authors:** Ilya Kister, Ryan Curtin, Jinglan Pei, Katherine Perdomo, Tamar E. Bacon, Iryna Voloshyna, Joseph Kim, Ethan Tardio, Yogambigai Velmurugu, Samantha Nyovanie, Andrea Valeria Calderon, Fatoumatta Dibba, Stanzin Idga, Marie I. Samanovic, Pranil Raut, Catarina Raposo, Jessica Priest, Mark Cabatingan, Ryan C. Winger, Mark J. Mulligan, Yury Patskovsky, Gregg J. Silverman, Michelle Krogsgaard

## Abstract

**Objective:** To compare ‘hybrid immunity’ (prior COVID-19 infection plus vaccination) and post-vaccination immunity to SARS CoV-2 in MS patients on different disease-modifying therapies (DMTs) and to assess the impact of vaccine product and race/ethnicity on post-vaccination immune responses.

**Methods:** Consecutive MS patients from NYU MS Care Center (New York, NY), aged 18-60, who completed COVID-19 vaccination series ≥6 weeks previously were evaluated for SARS CoV-2-specific antibody responses with electro-chemiluminescence and multiepitope bead-based immunoassays and, in a subset, live virus immunofluorescence-based microneutralization assay. SARS CoV-2-specific cellular responses were assessed with cellular stimulation TruCulture IFNγ and IL-2 assay and, in a subset, with IFNγ and IL-2 ELISpot assays. Multivariate analyses examined associations between immunologic responses and prior COVID-19 infection while controlling for age, sex, DMT at vaccination, time-to-vaccine, and vaccine product.

**Results:** Between 6/01/2021-11/11/2021, 370 MS patients were recruited (mean age 40.6 years; 76% female; 53% non-White; 22% with prior infection; common DMT classes: ocrelizumab 40%; natalizumab 15%, sphingosine-1-phosphate receptor modulators 13%; and no DMT 8%). Vaccine-to-collection time was 18.7 (±7.7) weeks and 95% of patients received mRNA vaccines. In multivariate analyses, patients with laboratory-confirmed prior COVID-19 infection had significantly increased antibody and cellular post-vaccination responses compared to those without prior infection. Vaccine product and DMT class were independent predictors of antibody and cellular responses, while race/ethnicity was not.

**Interpretation:** Prior COVID-19 infection is associated with enhanced antibody and cellular post-vaccine responses independent of DMT class and vaccine type. There were no differences in immune responses across race/ethnic groups.

## Introduction

Some of the disease-modifying therapies (DMTs) for MS suppress the immune system, which results in reduced immune responses to vaccinations ^1-25^. In a meta-analysis of COVID-19 vaccine studies in MS, post-vaccine seroconversion rates were 13-fold lower among patients on B-cell depleting anti-CD20 therapies (aCD20) and 8-fold lower with S1P receptor modulators (S1P) as compared to patients not on a DMT ^24^. Post-vaccine T-cell activation post-vaccine is suppressed with S1P ^18, 21, 22, 25^ but largely intact with B-cell depleting therapies even in the absence of antibody responses ^4, 6, 8, 10, 16-18, 21, 22, 25-27^.

An important unanswered question is whether post-vaccination immune responses are enhanced in MS patients who previously experienced SARS CoV-2 infection compared to those without prior infection. The clinical relevance of ‘hybrid immunity’ – infection plus vaccination - is increasing with the rising prevalence of SARS CoV-2 infection. As of November 2021 (pre-Omicron variant), 40% of the world population has already been infected with SARS CoV-2 at least once ^28^, and in some areas, the prevalence was even higher, e.g., 68% in South Africa ^29^. Since the advent of the Omicron variant, half of the US population was infected within months of this variant’s emergence ^30^. Thus, hybrid immunity is the dominant mode of immunity in most countries and, by extension, among the MS patients in these countries.

Several large, population-based studies have demonstrated that hybrid immunity affords a higher level of protection against reinfection and hospitalization than infection or vaccination alone ^31-35^. The main mechanism underlying enhanced immunity is “the strength and breadth of the antibody responses after vaccination of previously SARS-CoV-2–infected persons” with secondary contributions from Spike- and non-Spike-specific T cell memory ^36^. Thus, persons with prior infection (‘PI’) exhibit elevated induction of IFNγ-producing Spike-reactive CD4+ T cells following vaccination compared to those with no prior infection (‘NPI’) ^37)^ and expansion of spike-specific memory CD8+ T cells ^38^.

There is preliminary evidence that immunologic benefits of hybrid immunity may extend to patients with immune suppression, such as kidney and other solid organ transplant recipients. These patients have very attenuated humoral responses to vaccines but still exhibit a more robust post-vaccination antibody response if they had PI^39, 40^. How antibody and cellular responses to COVID-19 vaccines in MS patients on different DMTs compare in patients with and without prior COVID-19 infection has not, to our knowledge, been investigated. To address this question is the primary objective of our study. Additionally, we took advantage of our large and diverse dataset to investigate two other questions that have not received attention in the literature on post-vaccination responses in MS: 1. whether there is a difference in the immune response in MS patients between the two most commonly used mRNA vaccines - BNT162b2 (Pfizer-BioNTech) and mRNA-1273 (Moderna); and 2. whether post-vaccine immune responses vary by race/ethnicity (White, African-Americans, Hispanic-Americans, Other). The latter question is relevant in view of the assertion of “ample evidence that ethnicity affects responsiveness to vaccines” ^44^ on the one hand, and the paucity of data on vaccination responses among MS patients from underrepresented minorities on the other.

## Methods

### Study population

Consecutive patients who receive routine neurologic care at the NYU Multiple Sclerosis Comprehensive Care Center (New York City, NY) were invited to participate if they had clinician-diagnosed MS (revised 2017 McDonald criteria) ^45^; either treated with an FDA-approved DMT for MS or were not on any DMT; aged 18 to 60; had Expanded Disability Status Scale (EDSS) score of 0 (normal) to 7 (wheelchair-bound); and completed the FDA-approved COVID-19 vaccination series at least 6 weeks prior to blood sample collection. ‘Completed vaccination series’ was defined as two doses of Comirnaty (Pfizer-BioNTech), two doses of Spikevax (Moderna), or a single dose of adenoviral vector JNJ-78436735 (Johnson & Johnson) since this was the FDA-approved schedule for most the study period, i.e., before 3^rd^ dose/’vaccine boosters’ were recommended by CDC. Exclusion criteria were: concurrent immunosuppressive therapy; active systemic cancer; primary or acquired immunodeficiency; active drug or alcohol abuse; aCD20 therapy other than ocrelizumab (OCR); uncontrolled diabetes mellitus; end-organ failure (cardiac, pulmonary, renal, hepatic); systemic lupus erythematosus or other systemic autoimmune diseases. Patients were excluded if they received high-dose oral or parenteral corticosteroids, intravenous immunoglobulin (IVIG), plasmapheresis (PLEX), convalescent plasma, or polyclonal antibody treatments for COVID-19 within three months of sample collection; had COVID-19 symptom onset or tested positive by SARS-CoV-2 real-time PCR within two weeks of sample collection; received 3^rd^/booster dose of COVID-19 vaccines at any time before sample collection.

Enrollment period was from 6/01/2021 to 11/11/2021. All patients were interviewed by a trained research coordinator with a structured instrument. Patients were queried about: COVID-19 symptoms (per CDC clinical case definition ^46^; COVID-19 exposures from February 2020 to the time of enrollment; commercial SARS-CoV-2 testing dates and results (PCR or Antibody); COVID-19 treatments; COVID-19 vaccinations (vaccine product and dates); MS treatment at the time of vaccination. Electronic medical records were reviewed for COVID-19- and MS-relevant information. Laboratory confirmation required to confirm prior infection was either history of a positive test on SARS CoV-2 PCR, elevated antibodies to Nucleocapsid protein, or elevated anti-Spike antibodies prior to vaccination.

### Serological analyses

Patients’ serologic status was assessed using three different methods as described in our prior work ^42^. Briefly, we carried out the following tests:

1. Electro-chemiluminescence immunoassay using the Elecsys® platform (Roche Diagnostics GmbH, Mannheim, Germany), measuring antibodies to Nucleocapsid (N) (qualitative) and Receptor Binding Domain (RBD) of Spike (S) protein (quantitative). Values ≥1.0 U/mL were interpreted as ‘positive’ for anti-N SARS-CoV-2 antibodies and indicative of prior infection. For anti-Spike Abs, values of ⍰0.8 U/mL were considered ‘positive’, and those below the lower limit of quantification of the assay (<0.4 U/mL) were considered ‘negative’ and set to 0.4 U/mL ^47^. Any value above the upper limit of dilution of 1:25,000 was considered ‘positive’ and set to 25,000. Five samples (1%) exceeded the upper limit of dilution in our dataset.
2. NYU proprietary custom Multiepitope Bead-based Immunoassay (MBI) measures antibody responses to two recombinant proteins (Wuhan variant total Spike and receptor-binding domain (RBD) domain of Spike; Sino Biological cat no. 40590-V08B, 40592-V08B, 40591-V49H-B, respectively), using control analytes of Human serum albumin (HSA), tetanus toxoid and anti-human IgG (Jackson Immunoresearch, West Grove, PA, USA.) coupled to commercial paramagnetic beads (MagPix, Luminex) as previously described ^48, 49^. For reference, we provide MBI data for Spike and RBD for healthy, previously uninfected controls at baseline and three months post COVID-19 vaccine in **Figure S1**.
3. For a subset of available samples, we assessed the SARS-CoV-2 viral neutralization activity of plasma using an immunofluorescence-based assay that detects the neutralization of infectious virus (SARS-CoV-2 isolate USA-WA1/2020 (NR-52281, GenBank accession no. MT233526) in cultured Vero E6 cells (African Green Monkey Kidney; ATCC #CRL-1586). The methodology was described in detail in ^50^.

### Assays of SARS-CoV-2-specific T-cell response

In all patients, T-cell responses to SARS-CoV-2 Spike protein were assessed using TruCulture® stimulation system (Rules Based Medicine, Austin, TX, USA), in which whole blood samples are incubated for 48 hours at 37°C in the presence of whole Spike protein. Collected supernatants are analyzed by IFNγ and IL-2 cytokine quantitative assays (Thermo Fisher [Waltham, MA, USA]; Cat # ENEHIFNΓ and 50-112-5363, respectively). Samples were unavailable or failed quality assurance checks in 24 patients (6% of all patients) for TruCulture IFNγ and 23 patients (6%) for IL-2 assessment. In a subset of patients (n=40), we used ELISpot to corroborate results obtained with the TruCulture system. The methodology was described in detail in our prior publication ^51^.

### Statistical analyses

All patients were included in the analyses. Descriptive summaries of the results from the immunoassays were reported for continuous and categorical variables. Results that have heavily skewed distributions were normalized by log transformation (base 10). Mean, standard deviation (SD), median, and range were reported for continuous variables. For categorical variables, counts and percentage of patients with positive results were summarized. Correlation analyses were performed using the Spearman correlation. Comparisons of endpoints were performed between patients on the various DMTs and patients who were not on any DMT (‘no DMT’ reference group, n=30). We combined IFNβ, glatiramer acetate, teriflunomide, and fumarates into the ‘other DMT’ group as our preliminary analyses showed no differences in either humoral or antibody responses post-vaccine on these DMTs, while patients on OCR, S1P, natalizumab were analyzed separately because of differences in immune responses among these groups. For analyses of immune responses across race/ethnic groups, we compared patients who self-identified as White, African-American and Hispanic-American, and ‘Other’. Multivariate analyses were performed to compare immune responses in patients with PI and NPI while controlling for the following covariates: age; sex; vaccine product; DMT at time of vaccination (OCR vs. NTZ vs. S1P vs. other DMT vs. no DMT); and time from vaccination (last dose) to sample collection. Because S1P is known to affect the cellular responses, we conducted additional multivariate analyses that excluded S1P patients. Missing data were not imputed.

The study was approved by the Institutional Review Board of the NYU Grossman School of Medicine (New York).

## Results

### 1. Demographic and clinical characteristics of the patients

Demographic and clinical characteristics of 370 enrolled MS patients, their DMT at the time of vaccination, vaccine product and date(s) of administration, prior COVID-19 infection status, and COVID-19-relevant comorbidities are shown in **Table 1**. The patients were relatively young (mean [SD]), 40.6 [10.3] years, and mostly female (75.9%), with a disease duration of (mean [SD]) 11.9 [8.7] years. The majority of patients (53%) self-identified as non-White, which is consistent with the race/ethnic composition of NYU MS Care Center ^52^. The most common vaccine type was Pfizer-BioNTech (Comirnaty) - 217 (58.6%), followed by Moderna (Spikevax) - 134 (36.2%) and adenoviral vector/J&J in 19 (5.1%) patients. Time from completing vaccination series to sample collection was (mean [SD]), 18.7 [7.7]) weeks. Laboratory-confirmed SARS CoV-2 infection was documented in 82 patients (22%). Among patients with symptomatic COVID-19, the mean time from COVID-19 symptom onset to sample collection was 46.8 weeks (median [IQR], 42 [28.4, 69.9]). All infections occurred prior to vaccination; three patients were hospitalized for severe COVID.

**Table 1.** Demographic and clinical characteristics of patients with MS (N=370)

| Characteristic | All subjects<br>(N=370) |
| --- | --- |
| Age, years |  |
| Mean (SD) | 40.6 (10.3) |
| Median (Q1, Q3) | 41.0 (32.0, 49.0) |
| Female, n (%) | 281 (75.9) |
| Race/ethnicity, n (%) |  |
| White | 174 (47.0) |
| African American/Black | 73 (19.7) |
| Hispanic | 89 (24.1) |
| Other | 34 (9.2) |
| MS subtype, n (%) |  |
| RRMS (Relapsing Remitting) | 339 (91.6) |
| SPMS (Secondary Progressive) | 15 (4.1) |
| PPMS (Primary Progressive) | 12 (3.2) |
| PRMS (Progressive Relapsing) | 4 (1.1) |
| DMT at enrollment, n (%) |  |
| Ocrelizumab | 146 (39.5) |
| Natalizumab | 54 (14.6) |
| S1P | 48 (13.0) |
| Other DMT<sup>a</sup> | 92 (24.9) |
| No DMT | 30 (8.1) |
| Ambulatory status, n (%) |  |
| Fully ambulatory | 302 (81.6) |
| Impaired but no assistance | 24 (6.5) |
| Assistance with cane | 27 (7.3) |
| Assistance with walker | 13 (3.5) |
| Non-ambulatory wheelchair | 4 (1.1) |
| Prior COVID-19 infection, n (%)<sup>b</sup> |  |
| Yes | 82 (22.2) |
| No | 288 (77.8) |
| Vaccine, n (%) |  |
| Pfizer-BioNTech (Comirnaty) | 217 (58.6) |
| Moderna (Spikevax) | 134 (36.2) |
| Johnson & Johnson | 19 (5.1) |
| Time from last vaccine to collection, mean (SD), weeks | 18.7 (7.7) |
| Number of COVID-relevant comorbidities, n (%)<sup>c</sup> |  |
| 0 | 317 (85.7) |
| 1 | 47 (12.7) |
| 2 | 6 (1.6) |

DMT, disease-modifying therapy; MS, multiple sclerosis; Q, quartile; S1P, sphingosine-1-phosphate receptor modulators; SD, standard deviation.
<sup>a</sup> ‘Other DMTs’ included interferon- $\beta$ (n=7), glatiramer acetate (n=14), teriflunomide (n=13), and fumarates (n=58)
<sup>b</sup> All infections occurred prior to vaccination. Three patients were hospitalized for severe COVID-19 prior to vaccination: a man in early 50s with hypertension on OCR; a woman in early 40s with hypertension, diabetes mellitus, obesity on OCR; and an obese woman in early 30s who was on no DMT at the time of infection.
<sup>c</sup> ‘COVID-relevant comorbidities’ included hypertension, chronic obstructive pulmonary disease, cardiovascular disease, diabetes mellitus, obesity, sickle cell disease, chronic kidney disease, chronic liver disease, and (non-skin) cancer.

### 2. Post-vaccination antibody responses across DMTs in patients with and without prior COVID-19 infection

All patients underwent serologic testing by Elecsys for antibodies to Nucleocapsid and Spike RBD, and by MBI for whole Spike protein and RBD component of Spike. There was a strong correlation between whole Spike by MBI and Spike RBD by MBI (r=0.80, p<0.0001) and moderately strong between anti-RBD antibody levels by MBI and Elecsys (r=0.59, p<0.0001).

Anti-Spike antibody responses with these three assays stratified by DMT class in patients with PI and NPI are shown in **Figure 1**. Compared with patients on no DMT, patients on OCR and S1P exhibited significantly diminished antibody responses on all three assays, while patients on NTZ and other DMTs had similar antibody responses to the no DMT group. In the subset of patients with NPI, OCR and S1P were also associated with significantly lowered responses on all assays compared to patients on no DMT. The same pattern persisted within the subset of patients with PI, except that differences in antibody responses between S1P and no DMT did not reach statistical significance in two assays (there were only 12 patients with PI on S1P).

**Figure 1.**
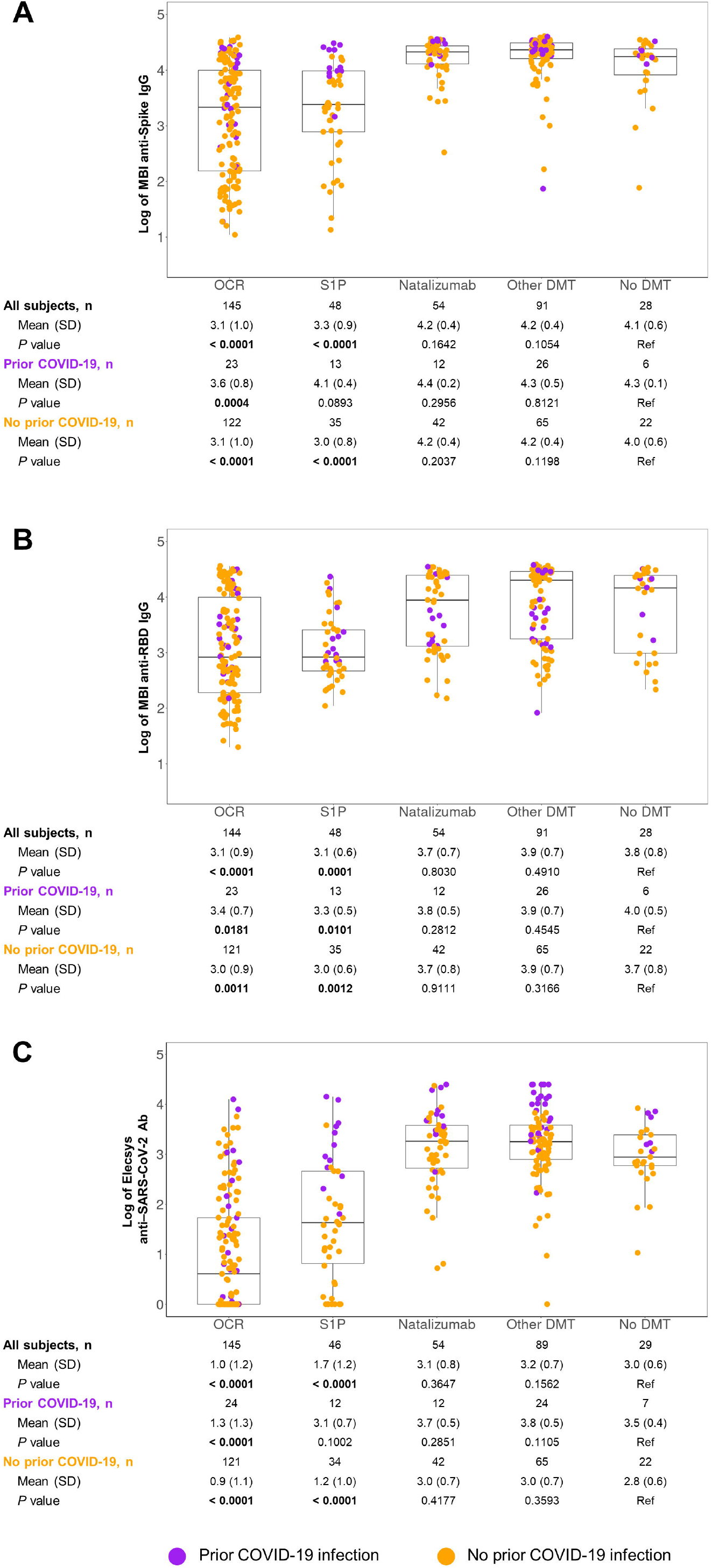
Post-vaccination antibody responses by DMT class^a^ and prior COVID-19 as assessed by **(A)** MBI anti-Spike IgG, **(B)** MBI anti-RBD Spike IgG, and **(C)** Elecsys anti-RBD Spike Ab Ab, antibody; DMT, disease-modifying therapy; IgG, immunoglobulin G; MBI, multiplex bead-based assay; RBD, receptor-binding domain. ^a^ ‘Other DMTs’ included interferon-b, glatiramer, fumarates, and teriflunomide. *P* values compare respective DMT classes vs no DMT (reference).

### 3. Functional neutralizing antibody (Nabs) in MS patients on different DMTs

Samples were available for testing functional neutralizing antibody (Nabs) titers via live virus microneutralization assay in 85 patients (23% of all enrollees) and these data is shown in **Table 2**. Nab levels showed a moderate correlation with anti-RBD antibody levels assessed by MBI assay (r=0.54, p<0.0001). Nab titers in 6 patients not on DMT were similar to patients on OCR and S1P, but lower than in the NTZ and ‘other DMT’ groups. Among those with PI (n=20), there were no differences by DMT, though the groups within this subset were very small: most had fewer than 5 patients, and there were no patient on S1P. Among patients with NPI (n=65), there were only 3 patients with no DMT (reference), and they had unexpectedly low Nabs titers, which may explain why patients in other groups (other than S1P) had relatively higher Nab titers. Across all DMTs, patients with PI had numerically higher Nabs titers than those with NPI.

**Table 2.** Neutralizing antibody titers by DMT class and prior COVID-19

| **Log of neutralizing antibodies-IC50** | **OCR** | **S1P** | **Natalizumab** | **Other DMT** | **No DMT (ref)** | **All subjects** |
| --- | --- | --- | --- | --- | --- | --- |
| All vaccinated subjects | N=146 | N=48 | N=54 | N=92 | N=30 | N=370 |
| n<sup>a</sup> | 42 | 6 | 14 | 17 | 6 | 85 |
| Mean (SD) | 1.4 (0.6) | 1.3 (0.4) | 1.7 (0.8) | 2.3 (0.9) | 1.3 (0.7) | 1.6 (0.8) |
| *P* value vs no DMT | 0.7669 | 0.8915 | 0.2718 | **0.0261** | ref | — |
| Prior COVID-19 | N=24 | N=13 | N=12 | N=26 | N=7 | N=82 |
| n<sup>a</sup> | 10 | 0 | 3 | 4 | 3 | 20 |
| Mean (SD) | 1.6 (0.6) | — | 2.3 (1.2) | 3.0 (0.4) | 1.6 (1.0) | 2.0 (0.9) |
| *P* value vs no DMT | 0.9490 | — | 0.4597 | 0.1340 | ref | — |
| No prior COVID-19 | N=122 | N=35 | N=42 | N=66 | N=23 | N=288 |
| n<sup>a</sup> | 32 | 6 | 11 | 13 | 3 | 65 |
| Mean (SD) | 1.3 (0.6) | 1.3 (0.4) | 1.6 (0.6) | 2.0 (0.8) | 1.0 (0.0) | 1.5 (0.7) |
| *P* value vs no DMT | **0.0046** | 0.2095 | **0.0120** | **0.0008** | ref | — |

DMT, disease-modifying therapy; IC50, half maximal inhibitory concentration.
<sup>a</sup> Number of patients tested for neutralizing antibodies.

### 4. Post-vaccination antibody responses stratified by COVID-19 mRNA vaccine product and race/ethnicity

Because the single-dose adenoviral vector vaccine is currently not recommended by the CDC ^53^ and because it was administered to only 5% of our patients, our analyses were focused on comparisons of immune responses to Pfizer-BioNTech (Comirnaty) and Moderna (Spikevax) only. The Moderna vaccine induced slightly higher antibody responses than Pfizer-BioNTech on whole Spike-MBI (mean [SD] 3.8 [0.9] vs. 3.6 [0.9] for Pfizer-BioNTech, p=0.013) and RBD Spike-MBI (3.6 [0.9] vs. 3.4 [0.8], p=0.02) assays, but the difference did not reach statistical significance on Elecsys assay (2.3 [1.5] vs. 2.0 [1.4], p=0.08). Among patients with PI, the two vaccines yielded similar immune responses across all assays (though always numerically larger for Moderna), while among NPI patients, Moderna induced higher antibody responses on whole Spike-MBI (mean [SD] 3.8 [0.9] for Moderna vs. 3.4 [1.0] for Pfizer-BioNTech, p=0.0026), RBD Spike-MBI (3.6 [0.9] vs. 3.3 [0.9], p=0.017), and RBD Spike Elecsys (mean [SD] 2.2 [1.4] vs. 1.7 [1.3], p=0.006). Antibody responses stratified by Pfizer-BioNTech and Moderna vaccines and DMT type are shown in **Figure 2**.

**Figure 2.**
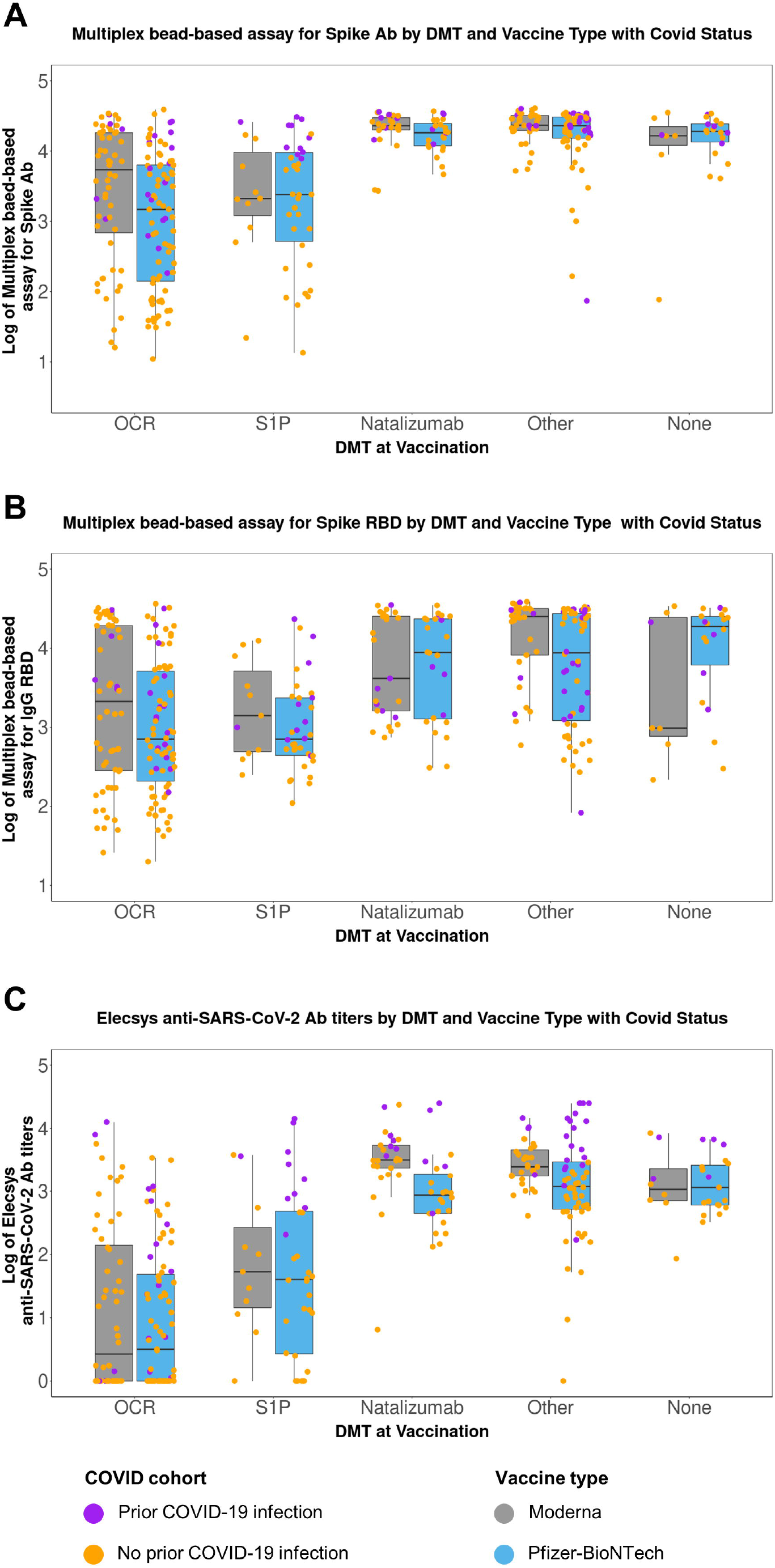
Post-vaccination antibody responses to Pfizer-BioNTech (Comirnaty) and Moderna (Spikevax) vaccine by DMT class^a^ and prior COVID-19 as assessed by **(A)** MBI anti-Spike IgG, **(B)** MBI anti-RBD IgG, and **(C)** Elecsys anti-SARS-CoV-2 Ab Ab, antibody; DMT, disease-modifying therapy; IgG, immunoglobulin G; MBI, multiplex bead-based assay; RBD, receptor-binding domain. ^a^ ‘Other DMTs’ included interferon-b, glatiramer, fumarates, and teriflunomide. *P* values compare respective DMT classes vs no DMT (reference).

SARS-CoV-2 antibody response across race/ethnicity groups (White vs. Black vs. Hispanics vs. Other), assessed with MBI and Elecsys and stratified by DMT, were similar as shown in **Figure 3A**.

**Figure 3.**
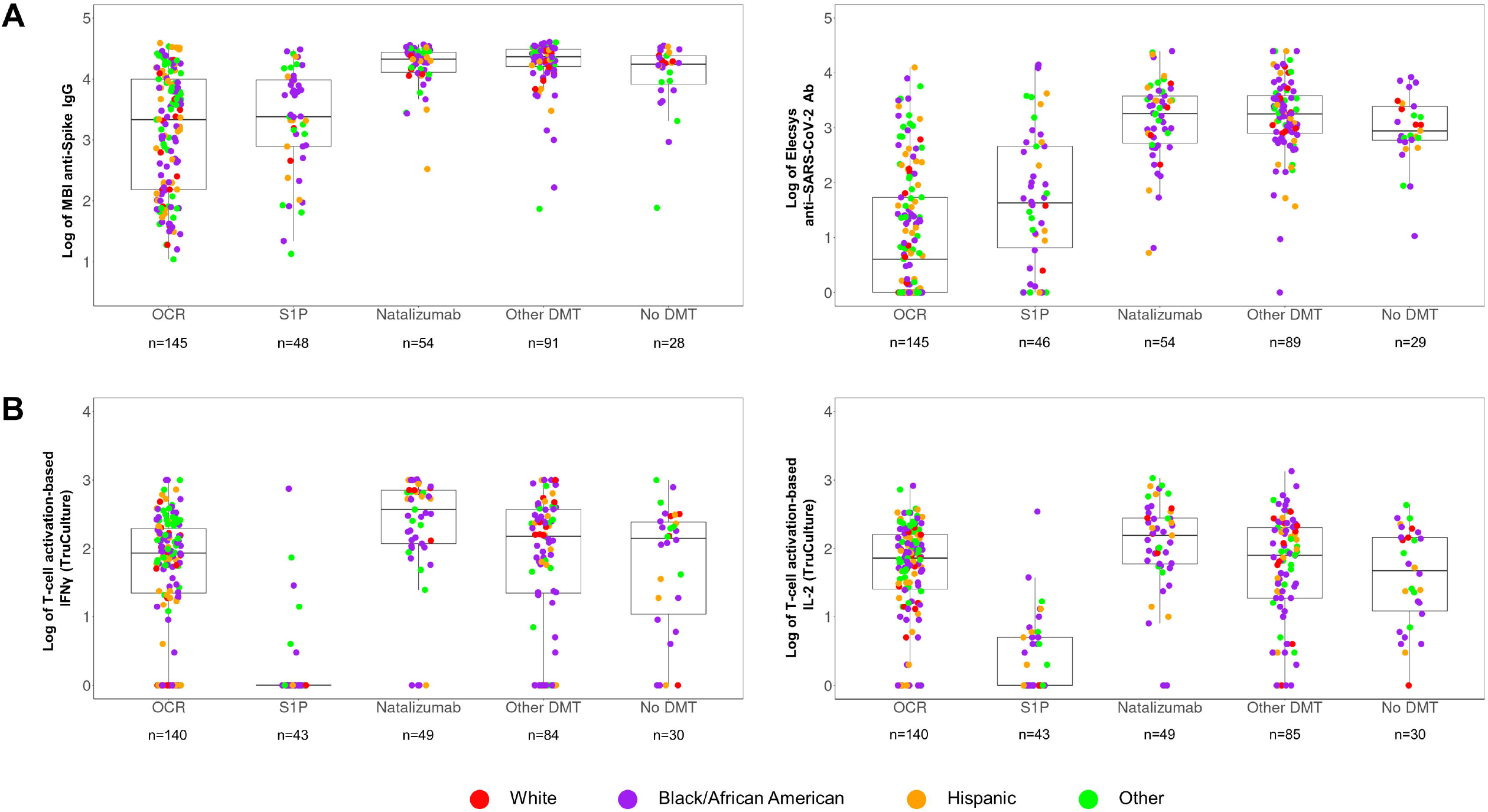
Post-vaccination **(A)** antibody responses as assessed by MBI anti-Spike and Elecsys anti-SARS-CoV-2 Ab by race and **(B)** cellular activation responses as assessed by TruCulture IFNγ and TruCulture IL-2 by race Ab, antibody; IFNγ, interferon gamma; IgG, immunoglobulin G; IL-2, interleukin 2; MBI, multiplex bead-based assay; RBD, receptor-binding domain.

### 5. Cellular activation responses to SARS CoV-2 antigens in patients on different DMTs with and without prior COVID-19 infection

Cellular activation TruCulture assay results stratified by DMT class are shown in **Figure 4A** (IFNγ) and **4B** (IL-2). Both IFNγ and IL-2 levels were depressed in S1P (p<0.0001) and increased in NTZ (p<0.01) relative to the ‘no DMT’, while patients on OCR and ‘other DMTs’ had responses similar to patients on no DMT. Comparing responses among DMTs by patients with PI and NPI, the patterns were similar: both PI and NPI patients on S1P had depressed levels of IFNγ and IL-2 from relative to respective PI/NPI patients on no DMT. For NTZ patients, IFNγ responses were significantly elevated for NPI patients, whereas PI patients only had increased IL-2 responses. Within each DMT class, including OCR, patients with PI had numerically higher or similar - but never lower - cellular activation responses compared to patients with NPI.

**Figure 4.**
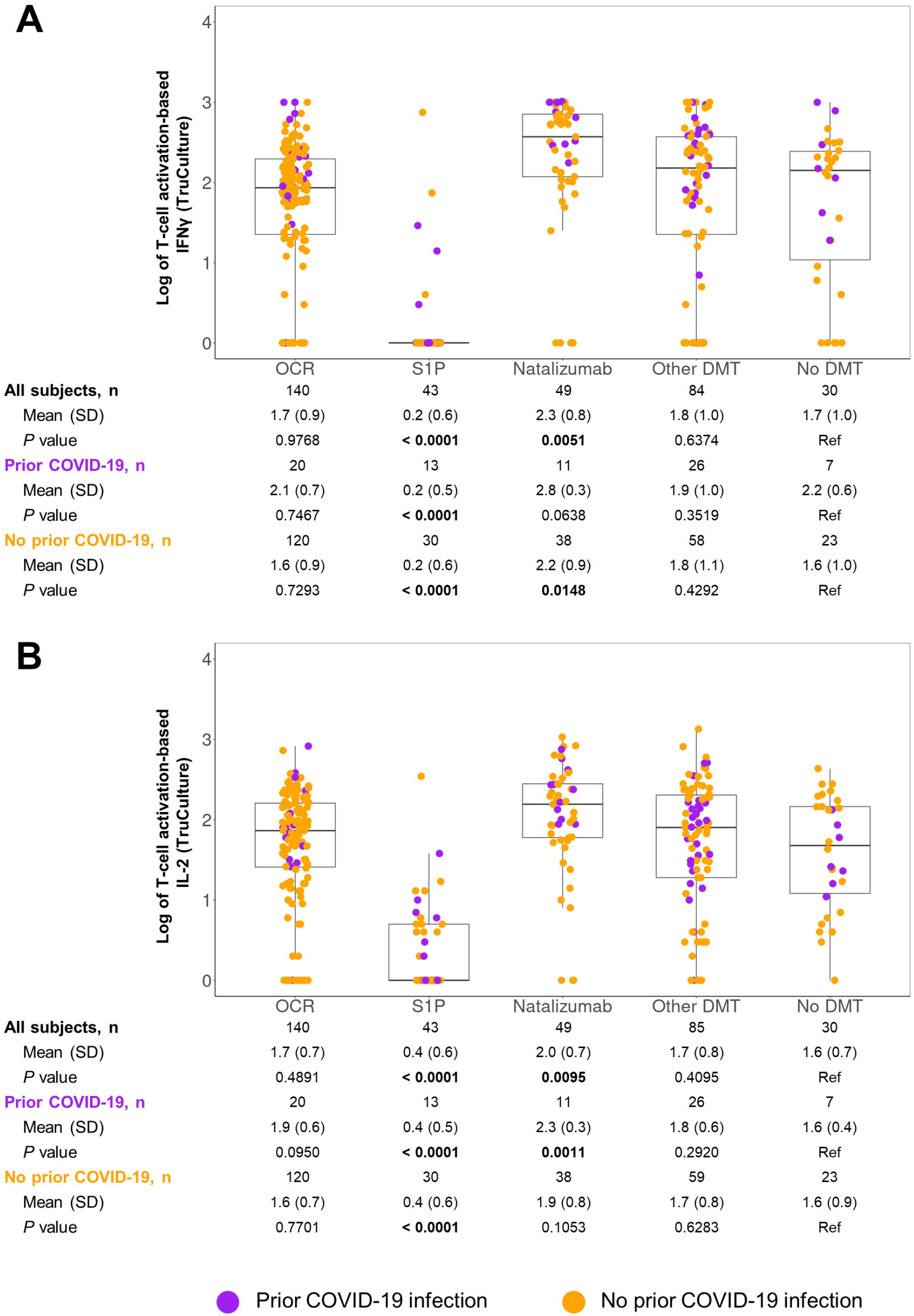
Post-vaccination cellular activation by DMT class^a^ and prior COVID-19 as assessed by **(A)** TruCulture IFNγ and **(B)** TruCulture IL-2 DMT, disease-modifying therapy; IFNγ, interferon gamma; IL-2, interleukin 2; SD, standard deviation. ^a^ ‘Other DMTs’ included interferon-b, glatiramer, fumarates, and teriflunomide. *P* values compare respective DMT classes vs no DMT (reference).

Lastly, we compared cellular activation in OCR patients with and without antibody response to the vaccine (as assessed with Elecsys). Cellular activation was nearly identical in the two subsets (**Figure S2**), suggesting that T-cell responses are largely independent of antibody responses.

### 6. Post-vaccination cellular responses stratified by COVID-19 mRNA vaccine and by race/ethnicity

Moderna vaccination resulted in slightly higher cellular responses than Pfizer-BioNTech for both TruCulture IFNγ (mean [SD] 1.9 [1.0] for Moderna and 1.5 [1.0] and for Pfizer-BioNTech; p=0.0006) and IL-2 (mean [SD] 1.8 [0.8] for Moderna and 1.5 [0.9] for Pfizer-BioNTech; p=0.0017) assays. Among patients with PI, Moderna induced statistically higher immune responses on IFNγ (mean [SD] 2.5 [0.5] vs. 1.6 [1.1] for Pfizer-BioNTech, p<0.0001) and IL-2 (2.0 [0.5] for Moderna, 1.6 [0.8] for Pfizer-BioNTech, p=0.02), Similarly among patients with NPI, Moderna induced statistically larger antibody responses on whole IFNγ (mean [SD] 1.5 [1.0] and 1.8 [1.0] for Pfizer-BioNTech and Moderna, respectively; p=0.012, and IL-2 (mean [SD] 1.5 [0.9] and 1.7 [0.8] for Pfizer-BioNTech and Moderna, respectively; p=0.007 assays. Cellular responses to Pfizer-BioNTech and Moderna vaccine stratified by DMT type are shown in **Figure 5**.

No differences in cellular activation by race/ethnicity were observed across DMTs as shown in **Figure 3B**.

**Figure 5.**
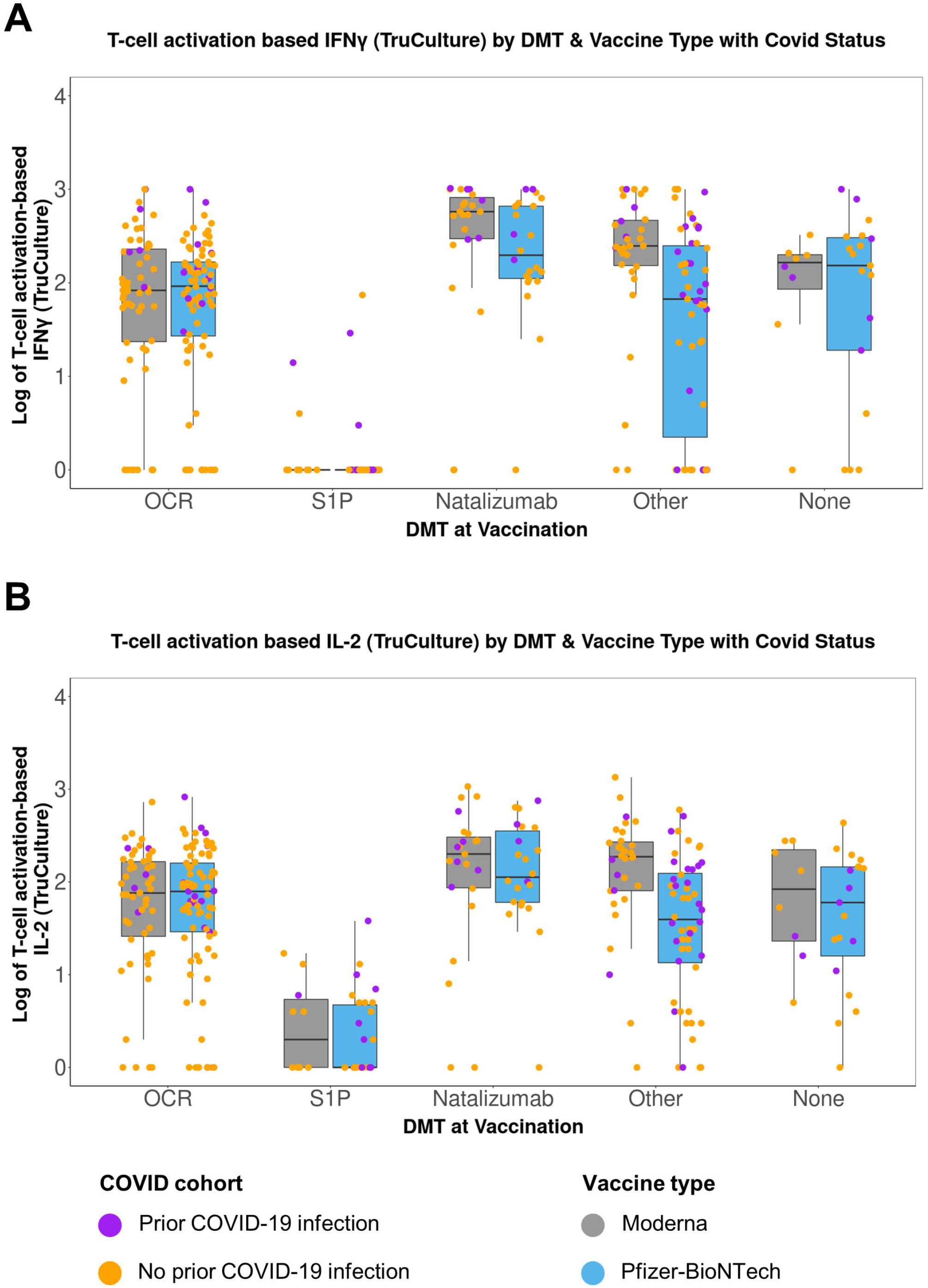
Post-vaccination T-cell activation responses to Pfizer-BioNTech (Comirnaty) and Moderna (Spikevax) vaccine by DMT class^a^ and prior COVID-19 as assessed by **(A)** TruCulture IFNγ and **(B)** TruCulture IL-2 DMT, disease-modifying therapy; IFNγ, interferon gamma; IL-2, interleukin 2; SD, standard deviation. ^a^ ‘Other DMTs’ included interferon-b, glatiramer, fumarates, and teriflunomide. *P* values compare respective DMT classes vs no DMT (reference).

### 7. Post-vaccination cellular responses assessed by ELISpot and stratified by DMT

T-cell responses for a subset of 40 patients were assessed using IFNγ and IL-2 ELISpot assays to corroborate the findings with TruCulture cell activation studies. The results are shown in **Figure S3**. Overall, the numerical trends with ELISpot across DMTs were similar to those seen with the TruCulture assay – T-cell activation was lower in S1P and higher for NTZ compared with no DMT group – but the comparisons did not reach statistical significance. This may be due to the fact that most patients in the ELISpot-tested subset were had PI (60%), for whom inter-DMT differences were less pronounced than among NPI, as well as to the small number of patients available for comparison (e.g., only 5 patients in the reference no DMT group). The correlation coefficient for TruCulture and ELISpot for IFNγ was r=0.33 (p=0.05) and borderline for IL-2, r=0.32 (p=0.06).

### 8. Multivariate comparison of antibody and cellular activation responses in PI and NPI patients

Multivariate analyses to examine associations between immunologic responses and COVID-19 status (PI vs. NPI) were carried out while controlling for age, sex, DMT class at the time of vaccination (OCR vs. S1P, vs. NTZ vs. other DMT, vs. no DMT), time-to-vaccine, and vaccine product (Pfizer-BioNTech vs. Moderna vs. J&J).

PI was a significant predictor of higher post-vaccine antibody responses in both Elecsys anti– SARS-CoV-2 Ab assay (antibody responses were 8.1-fold in PI vs. NPI, p<0.0001) and MBI Spike Ab assays (2.9-fold in PI, p<0.0001). Moreover, vaccine product and DMT class were highly significant predictors of antibody responses (p≤0.0001) for both assays and age was a significant predictor on Elecsys (p=0.0003) and MBI (p=0.026) assays. Time from vaccine-to-collection was weakly significant on MBI (p=0.044) but not on Elecsys (p=0.200) assay. Predictors of post-vaccine antibody are shown in **Table 3A**.

**Table 3.**
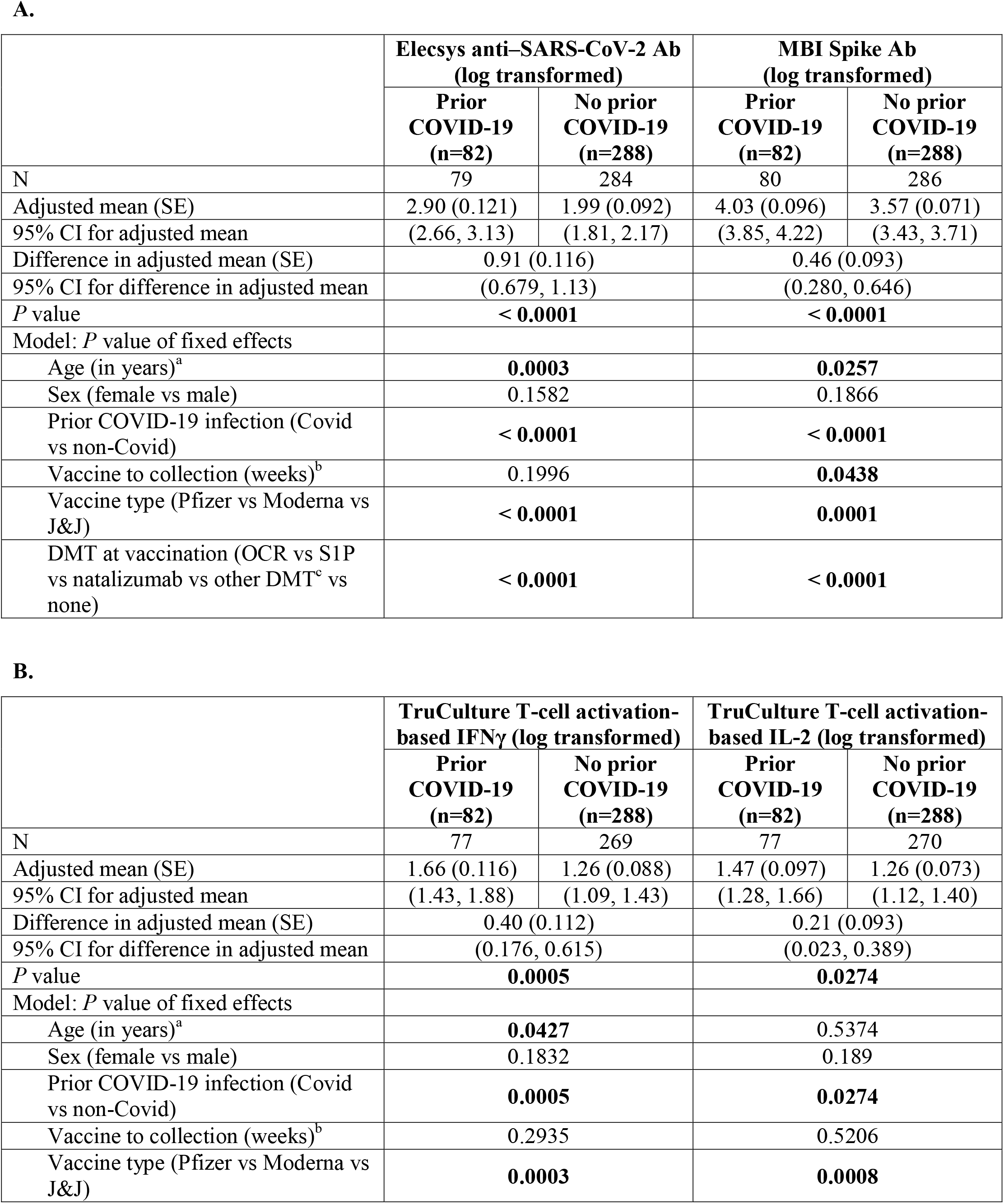
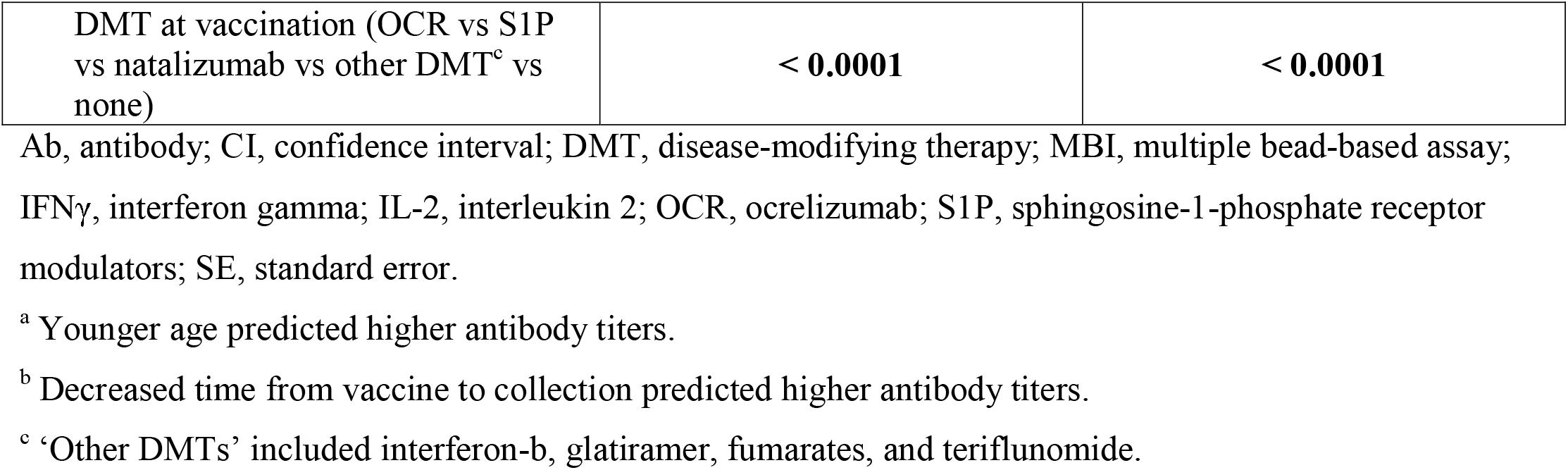
Differences between post-vaccination antibody responses **(A)** and cellular responses **(B)** in patients with and without prior COVID-19 infection and predictors of post-vaccination antibody in multivariate analyses

**A.**
|  | Elecsys anti-SARS-CoV-2 Ab<br>(log transformed) |  | MBI Spike Ab<br>(log transformed) |  |
| --- | --- | --- | --- | --- |
|  | Prior<br>COVID-19<br>(n=82) | No prior<br>COVID-19<br>(n=288) | Prior<br>COVID-19<br>(n=82) | No prior<br>COVID-19<br>(n=288) |
| N | 79 | 284 | 80 | 286 |
| Adjusted mean (SE) | 2.90 (0.121) | 1.99 (0.092) | 4.03 (0.096) | 3.57 (0.071) |
| 95% CI for adjusted mean | (2.66, 3.13) | (1.81, 2.17) | (3.85, 4.22) | (3.43, 3.71) |
| Difference in adjusted mean (SE) | 0.91 (0.116) |  | 0.46 (0.093) |  |
| 95% CI for difference in adjusted mean | (0.679, 1.13) |  | (0.280, 0.646) |  |
| *P* value | < 0.0001 |  | < 0.0001 |  |
| Model: *P* value of fixed effects |  |  |  |  |
| Age (in years)<sup>a</sup> | 0.0003 |  | 0.0257 |  |
| Sex (female vs male) | 0.1582 |  | 0.1866 |  |
| Prior COVID-19 infection (Covid vs non-Covid) | < 0.0001 |  | < 0.0001 |  |
| Vaccine to collection (weeks)<sup>b</sup> | 0.1996 |  | 0.0438 |  |
| Vaccine type (Pfizer vs Moderna vs J&J) | < 0.0001 |  | 0.0001 |  |
| DMT at vaccination (OCR vs S1P vs natalizumab vs other DMT<sup>c</sup> vs none) | < 0.0001 |  | < 0.0001 |  |

**B.**
| | TruCulture T-cell activation-based IFNγ (log transformed) | | TruCulture T-cell activation-based IL-2 (log transformed) | |
| --- | --- | --- | --- | --- |
|  | Prior<br>COVID-19<br>(n=82) | No prior<br>COVID-19<br>(n=288) | Prior<br>COVID-19<br>(n=82) | No prior<br>COVID-19<br>(n=288) |
| N | 77 | 269 | 77 | 270 |
| Adjusted mean (SE) | 1.66 (0.116) | 1.26 (0.088) | 1.47 (0.097) | 1.26 (0.073) |
| 95% CI for adjusted mean | (1.43, 1.88) | (1.09, 1.43) | (1.28, 1.66) | (1.12, 1.40) |
| Difference in adjusted mean (SE) | 0.40 (0.112) |  | 0.21 (0.093) |  |
| 95% CI for difference in adjusted mean | (0.176, 0.615) |  | (0.023, 0.389) |  |
| *P* value | 0.0005 |  | 0.0274 |  |
| Model: *P* value of fixed effects |  |  |  |  |
| Age (in years)<sup>a</sup> | 0.0427 |  | 0.5374 |  |
| Sex (female vs male) | 0.1832 |  | 0.189 |  |
| Prior COVID-19 infection (Covid vs non-Covid) | 0.0005 |  | 0.0274 |  |
| Vaccine to collection (weeks)<sup>b</sup> | 0.2935 |  | 0.5206 |  |
| Vaccine type (Pfizer vs Moderna vs J&J) | 0.0003 |  | 0.0008 |  |

|  |  |  |
| --- | --- | --- |
| DMT at vaccination (OCR vs S1P vs natalizumab vs other DMT<sup>c</sup> vs none) | **< 0.0001** | **< 0.0001** |

Ab, antibody; CI, confidence interval; DMT, disease-modifying therapy; MBI, multiple bead-based assay; IFNγ , interferon gamma; IL-2, interleukin 2; OCR, ocrelizumab; S1P, sphingosine-1-phosphate receptor modulators; SE, standard error.
<sup>a</sup> Younger age predicted higher antibody titers.
<sup>b</sup> Decreased time from vaccine to collection predicted higher antibody titers.
<sup>c</sup> ‘Other DMTs’ included interferon-b, glatiramer, fumarates, and teriflunomide.

PI was a consistent predictor of higher cellular activation responses as well. However, the magnitude of enhancement in PI was not as pronounced with respect to cellular responses as for antibody responses: 2.5-fold in PI vs. NPI on IFNγ assay (p<0.043) and 1.6-fold higher in PI vs. NPI on IL-2 assay (p=0.027). Vaccine type was a highly significant predictor of cellular activation based on induced IFNγ (p=0.0003) and IL-2 (p=0.008) levels, as was DMT class for both IFNγ and IL-2 (p<0.0001 for both). We assessed whether longer time to vaccine affected responses, but vaccine-to-collection time was not a predictor of cellular response on either assay. Age was a weakly significant predictor for IFNγ (p=0.043) but not for IL-2 (p=0.54). Predictors of post-vaccine cellular responses are summarized in **Table 3B**. As cellular responses in patients on S1P were markedly lower than those of other DMTs, we repeated multivariate analyses after excluding S1P patients. In these sensitivity analyses, prior infection, vaccine product, and DMT class all remained highly significant predictors of TruCulture IFNγ and IL-2 responses.

## Discussion

The literature on SARS CoV-2 vaccine responses in MS patients on various DMT is growing rapidly, but relatively little attention has been devoted to hybrid immunity. Most studies of post-vaccine responses in MS intentionally excluded patients with PI ^7, 18^ or had too few of them to allow for statistically meaningful comparison ^54, 55^. The studies that sought to account for the effect of PI reached contradictory conclusions: in two studies, enhanced humoral immune responses were seen in those with hybrid immunity ^15, 19^, and in one study, COVID-19 symptoms were not a predictor of immune response ^21^. In our study, enrollment of a large and representative sample of the MS clinic population (n=370), of whom 22% had laboratory-confirmed COVID-19 infections, enabled us to conduct multivariate comparisons of hybrid vs. vaccination-only immunity while accounting for relevant covariates – age, sex, DMT class, and time from vaccination to sample collection. We demonstrated that PI markedly enhances post-vaccine humoral and, and, to a lesser extent, cellular responses across DMTs, i.e. that the immunologic benefits of PI extended to patients on DMT classes - aCD20 and S1P - that are associated with attenuated post-infection ^41-43^ and post-vaccination responses ^24, 25^. Thus, vaccinated MS patients on aCD20 and S1P therapies who had prior COVID-19 infection - and these currently comprise the majority of patients in the US – likely have significantly better immune protection than would be expected based on published studies of vaccination-only immune responses ^24, 25, 56^. Moreover, our results suggest that it may be possible to improve humoral and cellular immune response defenses following repeated exposure to virus-specific antigens even in patients whose immune system has been partially compromised by medications. By its nature, exposure to viral infection is associated with unpredictable, potentially serious short- and long-term adverse events and is inadvisable for anyone, especially the immunocompromised. Additional vaccine dosing, on the other hand, is a time-tested, proven strategy. Whether the third dose of the COVID-19 vaccine is clinically effective in aCD20-treated MS patients has not been established. Several studies reported a minimal or very modest increase in levels of anti–SARS-CoV-2 following the third dose ^2, 11, 57^, especially in those without prior antibody response ^58^, while others have been more encouraging ^59^. The third dose of COVID-19 vaccine did boost T-cell responses in aCD20-treated patients, even those with undetectable memory response after the primary vaccine series ^54, 58^, but not in S1P-treated patients ^60^. We are currently conducting a longitudinal study to better understand the immunologic benefits of additional doses of COVID-19 vaccines in aCD20-treated patients (NCT04843774).

Another relevant question that has received relatively little attention is whether immune responses in MS patients on different DMTs vary by vaccine type. In the general population, two-dose mRNA-1273 (Moderna) appears to be slightly more efficacious than BNT162b2 (Pfizer-BioNTech) ^61, 62^. In MS patients, mRNA-1273 (Moderna) was associated with higher antibody titers than BNT162b2 (Pfizer-BioNTech) ^19^, but T-cell responses by vaccine product have not yet been investigated. In our study, Moderna vaccine induced higher antibody responses overall and, to some extent, enhanced cellular responses as well. Although it is unknown whether the minor immunologic differences in vaccine responses will translate into higher clinical effectiveness, e.g., better protection from serious infection, it is reasonable to recommend Moderna vaccine (when available), to patients who are receiving DMTs known to attenuate post-vaccination responses (especially if they had no known prior infection).

We leveraged the racial/ethnic diversity of our patient group – most of our patients self-identified as non-White – to investigate post-vaccination immune responses by race/ethnicity. There is a paucity of data on COVID-19 vaccine effectiveness in MS patients from underrepresented minorities, but there are reports that such responses in non-MS patients do differ by self-reported race/ethnicity. For example, African-Americans had enhanced antibody responses to an inactivated influenza vaccine and several other vaccines (reviewed in Kurpati et al. 2016), which may be due in part to higher proportion of peripheral CD19+ cells at baseline in persons of African ancestry ^63^. In our study, post-vaccine antibody or cellular responses were similar among Whites, African-Americans, and Hispanic Americans, regardless of whether they were treated with an aCD20 agent or not. This reassuring finding is consistent with what we observed with post-infection immune responses to SARS CoV-2 among unvaccinated patients ^42^ and reinforces the importance of vaccination across all race/ethnic groups.

An important strength of our study is the use of complementary techniques to assess both humoral responses (multiplex bead array, electro-chemiluminescence immunoassay, and, in a subset of patients, live microneutralization assay) and cellular responses (whole blood activation TruCulture IFNγ and Il-2 assays, and, in a subset of patients, post-stimulation ELISpot for IFNγ and Il-2), which allowed us to cross-validate our results. Reassuringly, the predictors of responses in multivariate analyses were the same for both MBI and Elecsys assay, except for time-to-vaccination, which was insignificant for Elecsys and significant for MBI (**Table 3A**). For cellular activation responses, the predictors were comparable for IFNγ and Il-2 assays, except for age, which was significant for IFNγ, but not for IL-2 (**Table 3B**). Interestingly, differences between PI and NPI were more pronounced for IFNγ than for IL-2. This may be due to a distinct population of IFNγ expressing memory SARS-CoV-2 spike-specific CD4+ T cells in patients with PI but not in those had vaccination only ^37^. Another strength of the study is relatively longer time from vaccination to sampling – a mean of ∼5 months – as compared to most other published studies in MS where patients were sampled within a few weeks of vaccination ^4, 6, 64, 65^. Our data provide reassurance on the durability of post-vaccination immune responses in treated MS patients, especially in those with prior infection.

A limitation of our study is the inability to definitively establish past infection in patients who did not undergo PCR testing at the time of symptoms or had an asymptomatic infection and did not have anti-spike antibody testing before vaccination. All of our patients were extensively queried about prior COVID-19 symptoms, exposures, and testing, and most patients had pre-vaccination antibody testing (including 90 patients who underwent such testing as part of our prior study ^42^). Nevertheless, there is a risk of misclassifying some PI patients as NPI exists - especially in aCD20 and S1P patients with attenuated post-infection humoral responses - and this could potentially skew the results towards the null hypothesis, i.e., artificially decrease the differences in immune responses between PI and NPI. Another limitation is that our patient population, while highly diverse and representative of our clinic with regard to both racial/ethnic composition and DMT usage, was relatively young (patients over age 60 were excluded) and therefore not very disabled. Our findings do not necessarily apply to older and more disabled patients. Lastly, it should be acknowledged that anti-Spike antibodies and cellular responses assessed in our study offer only a partial view of anti-SARS CoV-2 immunity. IgA antibodies, non-Spike antibody and T cell-function responses, memory B-cells, and innate lymphoid and related pathways likely also play roles in immunoprotection against SARS CoV-2. It remains to be determined to what extent immune differences between PI and NPI assessed with humoral and cellular responses to Spike antigen translate into clinical effectiveness.

In conclusion, our study demonstrated that the immunologic benefits of hybrid immunity extend to treated MS patients, including those on aCD20 and S1P therapies, and identified additional predictors of post-vaccination immune responses: vaccine product, DMT class, and, to a lesser extent, age. Longitudinal studies are needed to understand the interplay of waning immunity and re-exposures to SARS CoV-2 antigens through vaccination and infections among MS patients on different DMTs.

## Data Availability

All data produced in the present study are available upon reasonable request to the authors.

## Acknowledgments

This work was supported by an unrestricted investigator-initiated grant from Genentech. Editorial assistance for the figures and tables was provided by Health Interactions, Inc with the support of F. Hoffmann-La Roche Ltd. The authors are grateful to Drs Eric Deutsch and Erwan Muros for their critical review of the manuscript; to our medical assistants, Apphia Daniel and Teodora Drakulovic for their gentle bedside manner and great phlebotomy skills; and to all the patients who participated in this study.

## Author contributions

IK: conception and design of the study, acquisition and analysis of data, and drafting a significant portion of the manuscript or figures; RC: conception and design of the study, acquisition and analysis of data, and drafting a significant portion of the manuscript or figures; JP: conception and design of the study, acquisition and analysis of data, and drafting a significant portion of the manuscript or figures; KP: acquisition and analysis of data; TEB: acquisition and analysis of data; JK: acquisition and analysis of data; ET: acquisition and analysis of data; IY: acquisition and analysis of data; YV: acquisition and analysis of data; SN: acquisition and analysis of data; AVC: acquisition and analysis of data; FD: acquisition and analysis of data; SI: acquisition and analysis of data; MS: acquisition and analysis of data, and drafting a significant portion of the manuscript or figures; PR: acquisition and analysis of data; CR: conception and design of the study, acquisition and analysis of data; JP: conception and design of the study; MC: conception and design of the study; MJM: conception and design of the study, acquisition and analysis of data, and drafting a significant portion of the manuscript or figures; RCW: conception and design of the study; YP: conception and design of the study, acquisition and analysis of data, and drafting a significant portion of the manuscript or figures; GJS: conception and design of the study, acquisition and analysis of data, and drafting a significant portion of the manuscript or figures; MK: conception and design of the study, acquisition and analysis of data, and drafting a significant portion of the manuscript or figures.

## References

1. Achiron A, Mandel M, Dreyer-Alster S, et al. Humoral immune response to COVID-19 mRNA vaccine in patients with multiple sclerosis treated with high-efficacy disease-modifying therapies. Ther Adv Neurol Disord. 2021 04/22/2021;14:17562864211012835.

2. Achtnichts L, Jakopp B, Oberle M, et al. Humoral Immune Response after the Third SARS-CoV-2 mRNA Vaccination in CD20 Depleted People with Multiple Sclerosis. Vaccines (Basel). 2021 Dec 11;9(12).

3. Ali A, Dwyer D, Wu Q, et al. Characterization of humoral response to COVID mRNA vaccines in multiple sclerosis patients on disease modifying therapies. Vaccine. 2021 Oct 1;39(41):6111–6.

4. Apostolidis SA, Kakara M, Painter MM, et al. Cellular and humoral immune responses following SARS-CoV-2 mRNA vaccination in patients with multiple sclerosis on anti-CD20 therapy. Nat Med. 2021 Nov;27(11):1990–2001.

5. Bigaut K, Kremer L, Fabacher T, et al. Impact of Disease-Modifying Treatments of Multiple Sclerosis on Anti-SARS-CoV-2 Antibodies: An Observational Study. Neurol Neuroimmunol Neuroinflamm. 2021 Sep;8(5).

6. Brill L, Rechtman A, Zveik O, et al. Humoral and T-Cell Response to SARS-CoV-2 Vaccination in Patients With Multiple Sclerosis Treated With Ocrelizumab. JAMA Neurol. 2021 Dec 1;78(12):1510–4.

7. Capone F, Lucchini M, Ferraro E, et al. Immunogenicity and safety of mRNA COVID-19 vaccines in people with multiple sclerosis treated with different disease-modifying therapies. Neurotherapeutics. 2022 Jan;19(1):325–33.

8. Gadani SP, Reyes-Mantilla M, Jank L, et al. Discordant humoral and T cell immune responses to SARS-CoV-2 vaccination in people with multiple sclerosis on anti-CD20 therapy. EBioMedicine. 2021 Nov;73:103636.

9. Ghadiri F, Sahraian MA, Azimi A, Moghadasi AN. The study of COVID-19 infection following vaccination in patients with multiple sclerosis. Mult Scler Relat Disord. 2022 Jan;57:103363.

10. Katz JD, Bouley AJ, Jungquist RM, Douglas EA, O’Shea IL, Lathi ES. Humoral and T-cell responses to SARS-CoV-2 vaccination in multiple sclerosis patients treated with ocrelizumab. Mult Scler Relat Disord. 2022 Jan;57:103382.

11. Konig M, Lorentzen AR, Torgauten HM, et al. Humoral immunity to SARS-CoV-2 mRNA vaccination in multiple sclerosis: the relevance of time since last rituximab infusion and first experience from sporadic revaccinations. J Neurol Neurosurg Psychiatry. 2021 Oct 20.

12. Madelon N, Lauper K, Breville G, et al. Robust T cell responses in anti-CD20 treated patients following COVID-19 vaccination: a prospective cohort study. Clin Infect Dis. 2021 Nov 17.

13. Mariottini A, Bertozzi A, Marchi L, et al. Effect of disease-modifying treatments on antibody-mediated response to anti-COVID19 vaccination in people with multiple sclerosis. J Neurol. 2022 Jun;269(6):2840–7.

14. Ozakbas S, Baba C, Dogan Y, Cevik S, Ozcelik S, Kaya E. Comparison of SARS-CoV-2 antibody response after two doses of mRNA and inactivated vaccines in multiple sclerosis patients treated with disease-modifying therapies. Mult Scler Relat Disord. 2022 Feb;58:103486.

15. Pitzalis M, Idda ML, Lodde V, et al. Effect of Different Disease-Modifying Therapies on Humoral Response to BNT162b2 Vaccine in Sardinian Multiple Sclerosis Patients. Front Immunol. 2021 12/09/2021;12:781843.

16. Pompsch M, Fisenkci N, Horn PA, Kraemer M, Lindemann M. Evidence of extensive cellular immune response after SARS-CoV-2 vaccination in ocrelizumab-treated patients with multiple sclerosis. Neurol Res Pract. 2021 Nov 22;3(1):60.

17. Rauber S, Korsen M, Huntemann N, et al. Immune response to SARS-CoV-2 vaccination in relation to peripheral immune cell profiles among patients with multiple sclerosis receiving ocrelizumab. J Neurol Neurosurg Psychiatry. 2022 Feb 22.

18. Sabatino JJ, Jr., Mittl K, Rowles WM, et al. Multiple sclerosis therapies differentially affect SARS-CoV-2 vaccine-induced antibody and T cell immunity and function. JCI Insight. 2022 Feb 22;7(4).

19. Sormani MP, Inglese M, Schiavetti I, et al. Effect of SARS-CoV-2 mRNA vaccination in MS patients treated with disease modifying therapies. EBioMedicine. 2021 Oct;72:103581.

20. Stefanski AL, Rincon-Arevalo H, Schrezenmeier E, et al. B Cell Numbers Predict Humoral and Cellular Response Upon SARS-CoV-2 Vaccination Among Patients Treated With Rituximab. Arthritis Rheumatol. 2022 Jun;74(6):934–47.

21. Tallantyre EC, Vickaryous N, Anderson V, et al. COVID-19 Vaccine Response in People with Multiple Sclerosis. Ann Neurol. 2022 Jan;91(1):89–100.

22. Tortorella C, Aiello A, Gasperini C, et al. Humoral- and T-Cell-Specific Immune Responses to SARS-CoV-2 mRNA Vaccination in Patients With MS Using Different Disease-Modifying Therapies. Neurology. 2022 Feb 1;98(5):e541–e54.

23. Trumpelmann S, Schulte-Mecklenbeck A, Steinberg OV, et al. Impact of disease-modifying therapies on humoral and cellular immune-responses following SARS-CoV-2 vaccination in MS patients. Clin Transl Sci. 2022 Feb 25.

24. Etemadifar M ea. Multiple sclerosis disease-modifying therapies and COVID-19 vaccines: A practical review and meta-analysis. 2022; Available from: https://www.medrxiv.org/content/10.1101/2022.02.12.22270883v1.full.pdf.

25. Schietzel S, Anderegg M, Limacher A, et al. Humoral and cellular immune responses on SARS-CoV-2 vaccines in patients with anti-CD20 therapies: a systematic review and meta-analysis of 1342 patients. RMD Open. 2022 Feb;8(1).

26. Mrak D, Tobudic S, Koblischke M, et al. SARS-CoV-2 vaccination in rituximab-treated patients: B cells promote humoral immune responses in the presence of T-cell-mediated immunity. Ann Rheum Dis. 2021 Oct;80(10):1345–50.

27. Simon D, Tascilar K, Schmidt K, et al. Humoral and Cellular Immune Responses to SARS-CoV-2 Infection and Vaccination in Autoimmune Disease Patients With B Cell Depletion. Arthritis Rheumatol. 2022 Jan;74(1):33–7.

28. Barber RM, Sorensen RJD, Pigott DM, et al. Estimating global, regional, and national daily and cumulative infections with SARS-CoV-2 through Nov 14, 2021: a statistical analysis. The Lancet. 2022.

29. Madhi SA, Kwatra G, Myers JE, et al. Population Immunity and Covid-19 Severity with Omicron Variant in South Africa. N Engl J Med. 2022 Apr 7;386(14):1314–26.

30. Bedford. Continuing SARS-CoV-2 evolution under population immune pressure. 2022; Available from: https://www.fda.gov/media/157471/download.

31. Nordstrom P, Ballin M, Nordstrom A. Risk of SARS-CoV-2 reinfection and COVID-19 hospitalisation in individuals with natural and hybrid immunity: a retrospective, total population cohort study in Sweden. Lancet Infect Dis. 2022 Jun;22(6):781–90.

32. Plumb ID FL, Barkley E, et al. Effectiveness of COVID-19 mRNA Vaccination in Preventing COVID-19–Associated Hospitalization Among Adults with Previous SARS-CoV-2 Infection — United States, June 2021–February 2022 | MMWR. MMWR Morb Mortal Wkly Rep. 2022 2022-04-12T12:58:25Z.

33. Bates TA, McBride SK, Leier HC, et al. Vaccination before or after SARS-CoV-2 infection leads to robust humoral response and antibodies that effectively neutralize variants. Sci Immunol. 2022 Feb 18;7(68):eabn8014.

34. Suarez Castillo M, Khaoua H, Courtejoie N. Vaccine-induced and naturally-acquired protection against Omicron and Delta symptomatic infection and severe COVID-19 outcomes, France, December 2021 to January 2022. Euro Surveill. 2022 Apr;27(16).

35. Altarawneh HN, Chemaitelly H, Ayoub H, et al. Effect of prior infection, vaccination, and hybrid immunity against symptomatic BA.1 and BA.2 Omicron infections and severe COVID-19 in Qatar. 2022 2022-03-22.

36. Crotty S. Hybrid immunity. Science. 2021 2021/06/25/;372(6549):1392–3.

37. Rodda LB, Morawski PA, Pruner KB, et al. Imprinted SARS-CoV-2-specific memory lymphocytes define hybrid immunity. Cell. 2022 Apr 28;185(9):1588–601 e14.

38. Minervina AA, Pogorelyy MV, Kirk AM, et al. SARS-CoV-2 antigen exposure history shapes phenotypes and specificity of memory CD8(+) T cells. Nat Immunol. 2022 May;23(5):781–90.

39. Kemlin D, Lemy A, Pannus P, et al. Hybrid immunity to SARS-CoV-2 in kidney transplant recipients and hemodialysis patients. Am J Transplant. 2022 Mar;22(3):994–5.

40. Boyarsky BJ, Werbel WA, Avery RK, et al. Antibody Response to 2-Dose SARS-CoV-2 mRNA Vaccine Series in Solid Organ Transplant Recipients. JAMA. 2021 Jun 1;325(21):2204–6.

41. Asplund Hogelin K, Ruffin N, Pin E, et al. Development of humoral and cellular immunological memory against SARS-CoV-2 despite B cell depleting treatment in multiple sclerosis. iScience. 2021 Sep 24;24(9):103078.

42. Kister I, Patskovsky Y, Curtin R, et al. Cellular and Humoral Immunity to SARS-CoV-2 Infection in Multiple Sclerosis Patients on Ocrelizumab and Other Disease-Modifying Therapies: A Multi-Ethnic Observational Study. Ann Neurol. 2022 Jun;91(6):782–95.

43. Louapre C, Ibrahim M, Maillart E, et al. Anti-CD20 therapies decrease humoral immune response to SARS-CoV-2 in patients with multiple sclerosis or neuromyelitis optica spectrum disorders. J Neurol Neurosurg Psychiatry. 2022 Jan;93(1):24–31.

44. Kurupati R, Kossenkov A, Haut L, et al. Race-related differences in antibody responses to the inactivated influenza vaccine are linked to distinct pre-vaccination gene expression profiles in blood. Oncotarget. 2016 Sep 27;7(39):62898–911.

45. AJ T, BL B, F B, et al. Diagnosis of multiple sclerosis: 2017 revisions of the McDonald criteria. The Lancet Neurology. 2018 2018 Feb;17(2).

46. CDC Case Definition. [cited 2021 11/13/2021]; Available from: https://ndc.services.cdc.gov/case-definitions/coronavirus-disease-2019-2021/.

47. Jochum S, Kirste I, Hortsch S, et al. Clinical utility of Elecsys Anti-SARS-CoV-2 S assay in COVID-19 vaccination: An exploratory analysis of the mRNA-1273 phase 1 trial. medRxiv. 2021 Oct 19.

48. Pelzek AJ, Gronwall C, Rosenthal P, et al. Persistence of Disease-Associated Anti-Citrullinated Protein Antibody-Expressing Memory B Cells in Rheumatoid Arthritis in Clinical Remission. Arthritis Rheumatol. 2017 Jun;69(6):1176–86.

49. Radke EE, Brown SM, Pelzek AJ, et al. Hierarchy of human IgG recognition within the Staphylococcus aureus immunome. Sci Rep. 2018 Sep 5;8(1):13296.

50. Samanovic MI, Cornelius AR, Gray-Gaillard SL, et al. Robust immune responses are observed after one dose of BNT162b2 mRNA vaccine dose in SARS-CoV-2 experienced individuals. Sci Transl Med. 2021:eabi8961.

51. Kister I, Patskovsky Y, Curtin R, et al. Cellular and humoral immunity to SARS-CoV-2 infection in multiple sclerosis patients on ocrelizumab and other disease-modifying therapies: a multi-ethnic observational study. 2022 2022-01-10.

52. Kister I, Bacon T, Cutter GR. How Multiple Sclerosis Symptoms Vary by Age, Sex, and Race/Ethnicity. Neurol Clin Pract. 2021 Aug;11(4):335–41.

53. http://@CDCgov. CDC Endorses ACIP’s Updated COVID-19 Vaccine Recommendations | CDC Online Newsroom | CDC. 2021 2021-12-17T02:25:43Z.

54. Madelon N, Heikkila N, Sabater Royo I, et al. Omicron-Specific Cytotoxic T-Cell Responses After a Third Dose of mRNA COVID-19 Vaccine Among Patients With Multiple Sclerosis Treated With Ocrelizumab. JAMA Neurol. 2022 Apr 1;79(4):399–404.

55. Madelon N, Lauper K, Breville G, et al. Patients treated with anti-CD20 therapy can mount robust T cell responses to mRNA-based COVID-19 vaccines 2021 2021/07/23/.

56. Vijenthira A, Gong I, Betschel SD, Cheung M, Hicks LK. Vaccine response following anti-CD20 therapy: a systematic review and meta-analysis of 905 patients. Blood Adv. 2021 Jun 21;5(12):2624–43.

57. Achiron A, Mandel M, Gurevich M, et al. Immune response to the third COVID-19 vaccine dose is related to lymphocyte count in multiple sclerosis patients treated with fingolimod. J Neurol. 2022 May;269(5):2286–92.

58. Brill L, Raposo C, Rechtman A, et al. Severe Acute Respiratory Syndrome Coronavirus 2 Third Vaccine Immune Response in Multiple Sclerosis Patients Treated with Ocrelizumab. Ann Neurol. 2022 Jun;91(6):796–800.

59. Maglione A, Morra M, Meroni R, Matta M, Clerico M, Rolla S. Humoral response after the booster dose of anti-SARS-CoV-2 vaccine in multiple sclerosis patients treated with high-efficacy therapies. Mult Scler Relat Disord. 2022 May;61:103776.

60. Palomares Cabeza V, Kummer LYL, Wieske L, et al. Longitudinal T-Cell Responses After a Third SARS-CoV-2 Vaccination in Patients With Multiple Sclerosis on Ocrelizumab or Fingolimod. Neurol Neuroimmunol Neuroinflamm. 2022 Jul;9(4).

61. http://@CDCgov. Comparative Effectiveness of Moderna, Pfizer-BioNTech, and Janssen (Johnson & Johnson) Vaccines in Preventing COVID-19 Hospitalizations Among Adults Without Immunocompromising Conditions — United States, March–August 2021 | MMWR. 2021 2021-09-22T01:24:47Z.

62. Dickerman BA, Gerlovin H, Madenci AL, et al. Comparative Effectiveness of BNT162b2 and mRNA-1273 Vaccines in U.S. Veterans. N Engl J Med. 2022 Jan 13;386(2):105–15.

63. Tollerud DJ, Clark JW, Brown LM, et al. The influence of age, race, and gender on peripheral blood mononuclear-cell subsets in healthy nonsmokers. J Clin Immunol. 1989 May;9(3):214–22.

64. Gadani SP, Reyes-Mantilla M, Jank L, et al. Discordant humoral and T cell immune responses to SARS-CoV-2 vaccination in people with multiple sclerosis on anti-CD20 therapy 2021 2021/08/25/.

65. Sabatino JJ, Mittl K, Rowles W, et al. Impact of multiple sclerosis disease-modifying therapies on SARS-CoV-2 vaccine-induced antibody and T cell immunity 2021 2021/09/20/.

