## Supplemental Figures 1-3 for "Hybrid and vaccine-induced immunity against SARS-CoV-2 in MS patients on different disease-modifying therapies"

**Supplemental Figure 1.** Post-vaccination antibody responses in healthy, previously unaffected control subjects at baseline and 3 months post-COVID-19 vaccine as assessed by MBI anti-Spike

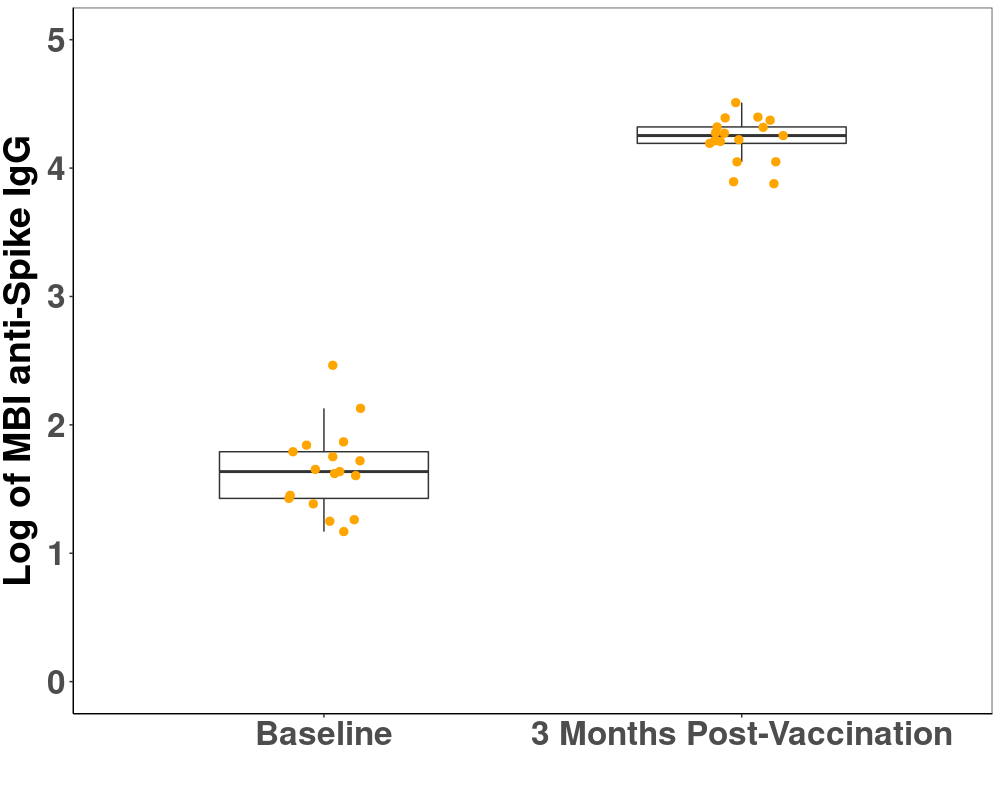

IgG, immunoglobulin G; MBI, multiplex bead-based assay.

Ab, antibody; IgG, immunoglobulin G; MBI, multiplex bead-based assay; RBD, receptor-binding domain.

**Supplemental Figure 2.** Comparison of post-vaccination T-cell activation in OCR patients with detectable and undetectable anti-Spike antibody response (assessed with Elecsys)

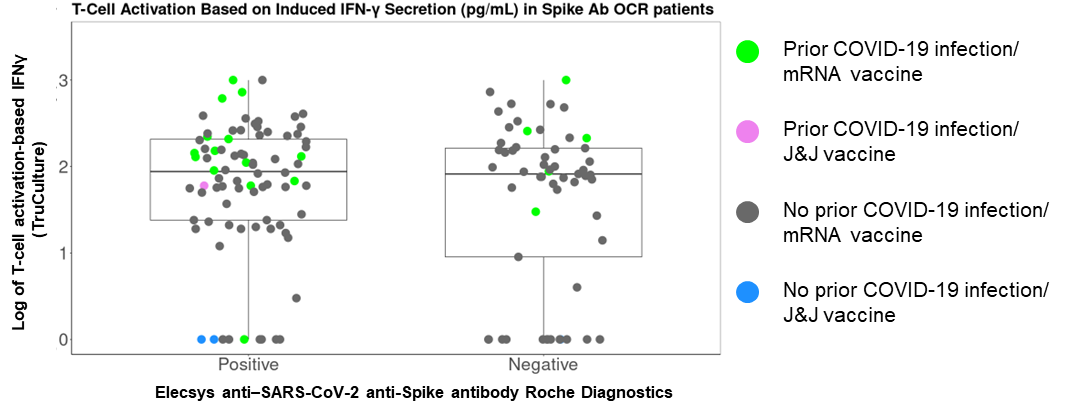

IFNγ, interferon gamma; OCR, ocrelizumab.

**Supplemental Figure 3.** Post-vaccination T-cell activation by DMT class^a^ and prior COVID-19 as assessed by **(A)** ELISpot IFNγ and **(B)** ELISpot IL-2

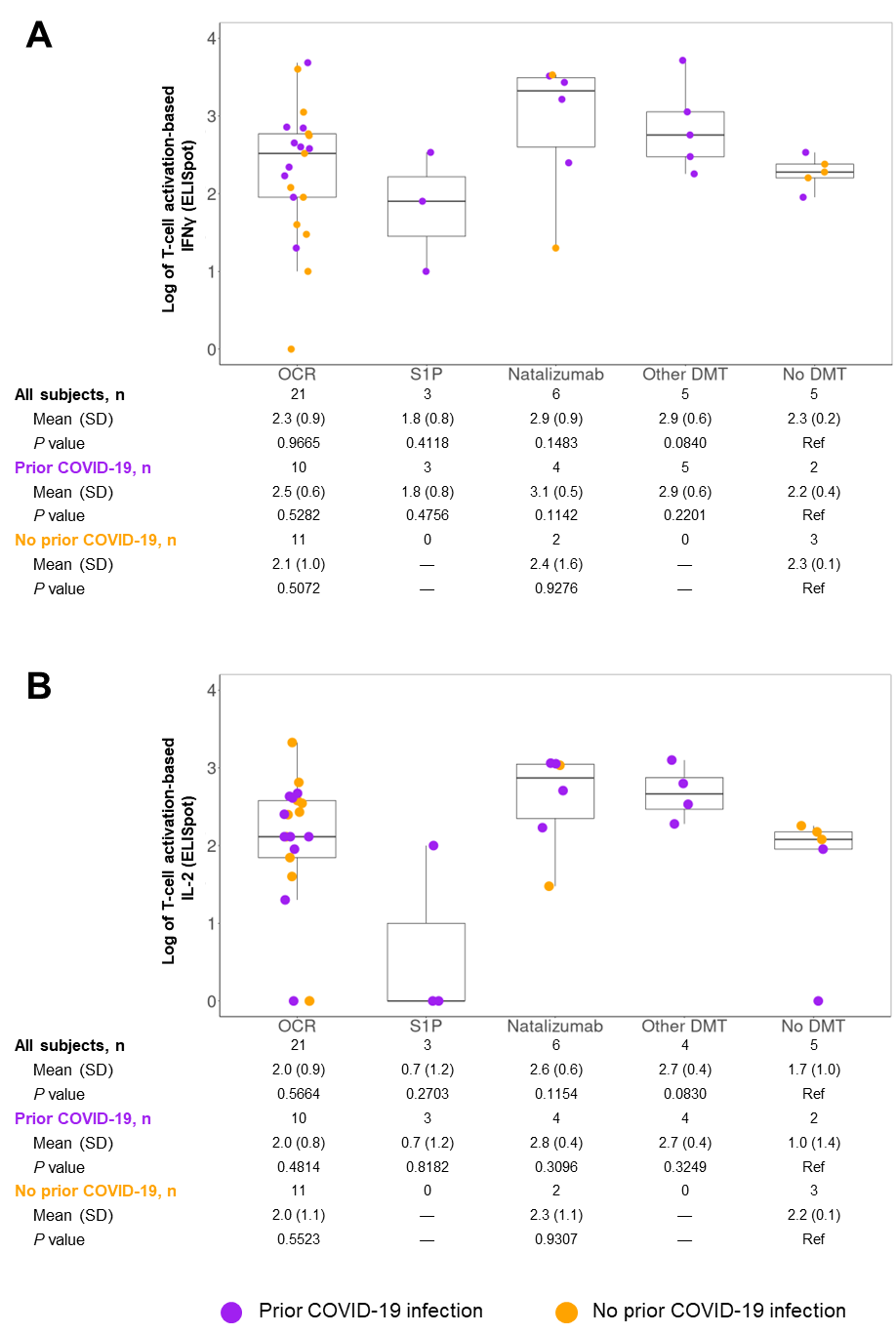

DMT, disease-modifying therapy; IFNγ, interferon gamma; IL-2, interleukin 2; SD, standard deviation.

^a^ ‘Other DMTs’ included interferon-b, glatiramer, fumarates, and teriflunomide. *P* values compare respective DMT classes vs no DMT (reference).
